# Beyond sex differences: equivalent adaptations across the O_2_ transport chain after exercise-based cardiac rehabilitation in women and men with coronary heart disease

**DOI:** 10.64898/2026.05.20.26353671

**Authors:** Mathieu Gayda, Florent Besnier, Pierre-Marie Leprêtre, Lukas-Daniel Trachsel, Josep Iglesies-Grau, Maxime Boidin, Pierre-Olivier Magnan, Damien Vitiello, Marine Kirsch, Axel Girault, Julie Lalongé, Martin Juneau, Anil Nigam, Louis Bherer

## Abstract

**Background:** Exercise-based cardiac rehabilitation (CR) improves peak oxygen uptake (V̇O_2_peak) in patients with coronary heart disease (CHD); however, whether women and men exhibit similar adaptations across the steps of O_2_ transport remains unknown. We aimed to compare the ventilatory and circulatory determinants of V̇O_2_peak changes between women and men with CHD following a structured exercise training program.

**Methods:** A total of 28 women (27%) and 75 men (73%) with CHD, matched for age, body mass index, and V̇O_2_peak (% predicted), underwent maximal cardiopulmonary exercise testing (CPET) before and after 12 weeks of CR. V̇O_2_peak and minute ventilation (V̇E) were measured breath by breath. Heart rate and cardiac output (Q̇c)were assessed non-invasively using impedance cardiography. Exercise efficiency (ΔV̇O_2_/ΔW), alveolar ventilation (V̇A), ventilatory efficiency (OUES), O_2_ pulse, arteriovenous oxygen content difference (C(a-v̄)O_2_) and gross muscular efficiency (ηW) were calculated using standard equations. Mixed model analyses (sex∈time) were used to compare training-induced changes between sexes.

**Results:** At baseline, values of V̇O_2_peak (absolute and normalized by fat free mass), V̇E, V̇A, O_2_ pulse, C(a-v̄)O_2_, ΔV̇O_2_/ΔW, ηW were significantly lower in women than in men with CHD (group effect, p<0.01). V̇O_2_peak normalized by fat-free mass improved similarly in both sexes after CR (p<0.0001, no significant sex x time interaction). Pulmonary convection (V̇E, V̇A), ventilatory efficiency (OUES), circulatory convection (Q̇c, cardiac index, O_2_ pulse), and peripheral gross muscular efficiency (ηW) all improved similarly after CR in women and men (effect size∈time effect, p<0.05, no significant group∈time interaction). The prevalence of responder categories did not differ between sexes (p=0.826).

**Conclusion:** Women and men with CHD demonstrated equivalent O_2_ transport phenotype adaptations after CR, with comparable improvements across the O_2_ transport chain (pulmonary, circulatory, and peripheral determinants of V̇O_2_peak).

## Introduction

Exercise-based cardiac rehabilitation program (CR) is a class I recommendation in individuals with coronary heart disease (CHD). CR confers long-term benefits, including reductions in cardiovascular events and mortality, alongside improvements in quality of life, cardiorespiratory fitness, and CHD risk factors ^1–3^. Despite being strongly recommended for both sexes, women with CHD remain underrepresented in CR programs, with referrals being less frequent ^4^ and exercise-based clinical trials including only about 17% women participants^1^. Cardiorespiratory fitness (measured by peak oxygen uptake: V̇O_2_peak), is a key prognostic marker in women and men with CHD, with its improvement linked to reduced mortality and cardiovascular events ^5–8^. Literature reports conflicting findings ^9^ on V̇O_2_peak improvements after CR in women and men with CHD, with some studies showing greater gains in men ^10–17^, while others report similar improvements between sexes ^18–25^. These discrepancies may partly reflect methodological heterogeneity, including differences in baseline matching, V̇O_2_peak assessment (directly measured vs. estimated), normalization methods, and training characteristics across studies. Beyond these differences, V̇O_2_peak is determined by several factors across the steps of O_2_ transport, including ventilatory, pulmonary gas exchange, cardiac, and peripheral muscular levels ^26, 27^. Convective O_2_ transport includes both pulmonary airflow and circulatory blood flow. Consequently, the pulmonary convective response to peak exercise can be characterized by parameters such as minute ventilation (V̇E) and alveolar ventilation (V̇A) ^26, 27^. Circulatory convection can be assessed during cardiopulmonary exercise testing (CPET) through O_2_ pulse, and, when cardiac output is measured, by indices such as peak cardiac output (Q̇c), peak cardiac index (CI), and peak cardiac power (CP) ^26, 28^. O_2_ diffusion refers to the transfer of O_2_ from the pulmonary capillaries into the blood circulation (pulmonary diffusion) and from the muscle capillaries into the mitochondria (peripheral diffusion) ^26, 27^. Pulmonary diffusion can be assessed by CPET index, such as the alveolar capillary gradient in O_2_ (PAi-aO_2_), end-tidal pressures of O_2_ (PETO_2_) and CO_2_ (PETCO_2_) ^26, 27^. The arteriovenous O_2_ difference (C(a-v̅)O_2_), the slope of O_2_ uptake relative to work rate change (ΔV̇O_2_/ΔWatts) and gross efficiency of external work (ηW) reflect peripheral O_2_ diffusion as well as the metabolic efficiency of the musculoskeletal system in converting chemical energy into mechanical work ^26, 29^. Women and men have different organ sizes and physiological functions across the steps of O_2_ transport that might influence their response to aerobic training ^30, 31^. In particular, women have lower absolute pulmonary V̇E and diffusing capacity, as well as smaller ventricular dimensions and Q̇c during peak exercise ^30, 31^. However, women show a similar pulmonary gas exchange vs. men ^32^ and a higher O_2_ extraction by muscle, higher mechanical efficiency and vascular compliance ^30, 33^ . It thus seems that in a healthy population, sex-specific differences exist in peripheral vs. central adaptations to exercise. To our knowledge, this has never been studied in women and men with CHD. Accordingly, the objective of this study was to compare the ventilatory and circulatory determinants of the underlying changes in peak oxygen uptake (ΔV̇O_2_peak) between women and men with CHD after an exercise-based CR program. Based on previous studies mentioned above, we hypothesized that ΔV̇O_2_peak will rely more on peripheral improvements after CR in women than in men.

## Material and Methods

### Study Structure and Patient Description

The baseline clinical characteristics of the patients are shown in **Table 1**. All patients were directed to a multi-disciplinary secondary prevention program at the Preventive medicine and physical activity of the Montreal Heart Institute and enrolled in a randomized exercise training intervention study ^22^. This study is based on a pooled analysis of data from four prospective randomized controlled trials on exercise training interventions in patients with CHD. All study protocols were approved by the Research Ethics and New Technology Development Committee of the Montreal Heart Institute and registered with ClinicalTrials.gov (Nos. NCT03414996, NCT02048696, NCT03443193, NCT04971707). Each patient provided written informed consent. All patients with CHD received optimal medical therapy following coronary revascularization. The inclusion and exclusion criteria for patients with CHD are detailed in the supplementary materials.

**Table 1.** Baseline characteristics of women and men with coronary heart disease (CHD).

| **Variable** | **Women, n=28**<br>**(mean±SD)** | **Men, n=75**<br>**(mean±SD)** | ***p*-value** |
| --- | --- | --- | --- |
| Age (years) | 65.5±5.9 | 66.4±5.7 | 0.498 |
| Height (cm) | 159.2±7.5 | 173.2±6.3 | <0.001 |
| Body mass (kg) | 73.8±16.5 | 84.7±15.2 | <0.001 |
| LBM (kg) | 44.2±6.6 | 62.0±8.1 | <0.001 |
| Body mass index (kg/m<sup>2</sup>) | 29.1±5.8 | 28.3±4.9 | 0.533 |
| Fat mass (%) | 39.6±5.4 | 26.1±5.9 | <0.001 |
| VO<sub>2</sub> peak (in % predicted) | 98.9±16.2 | 93.4±17.5 | 0.079 |
| **Peri-procedural characteristics** |  |  |  |
| Post-ACS | 21 (75) | 67 (89) | 0.067 |
| Recent ACS (<6weeks) | 20 (71) | 51 (68) | 0.806 |
| STEMI | 6 (21) | 25 (33) | 0.241 |
| PCI | 28 (100) | 65 (86) | 0.042 |
| CABG | 0 (0) | 8 (10) | 0.072 |
| **Cardiovascular risk profile** |  |  |  |
| Active smoking | 2 (7) | 5 (6) | 0.932 |
| Hypertension | 16 (57) | 47 (62) | 0.609 |
| Dyslipidemia | 22 (78) | 60 (80) | 0.873 |
| Type 2 diabetes mellitus | 4 (14) | 5 (6) | 0.201 |
| Obesity | 15 (53) | 26 (34) | 0.081 |
| Family history CVD | 8 (28) | 21 (28) | 0.954 |
| **Baseline medication** |  |  |  |
| Aspirin | 18 (64) | 44 (58) | 0.604 |
| Other APT | 17 (60) | 35 (46) | 0.205 |
| Lipid-lowering therapy | 21 (75) | 66 (88) | 0.198 |
| RAAS inhibitors | 15 (53) | 38 (50) | 0.793 |
| Beta-blockers | 21 (75) | 50 (66) | 0.416 |
| CCB | 1 (3) | 10 (13) | 0.154 |
| Diuretics | 1 (3) | 2 (2) | 0.808 |
| Nitrates | 7 (25) | 17 (22) | 0.803 |

Data are expressed as mean ± SD for parametric data or as median [interquartile range] for non-parametric data, dichotomous variables are expressed as numbers and percentages. ACS, acute coronary syndrome; APT, antiplatelet therapy; C, cholesterol; CABG, coronary-artery bypass grafting; CCB, calcium channel blocker; CVD, cardiovascular disease; HbA1c, glycosylated hemoglobin; LBM, lean body mass; PCI, percutaneous coronary intervention; RAAS inhibitors, inhibitor of the renin angiotensin aldosterone system; STEMI, ST elevation myocardial infarction.

### Assessment

Baseline clinical assessment, including medical history, physical examination, anthropometric measurements, and body composition analysis (bioimpedance, BC418 model; Tanita, Japan), was conducted at the start and upon completion of the program.

### Cardiopulmonary exercise testing

All patients underwent a maximal CPET on a cycle ergometer (Ergoline 800S, Bitz, Germany), following the guidelines of the American Heart Association and as previously described ^22, 34–36^. After a 3-min warm-up at 20 Watts (W), mechanical power was increased by 10–15 W per minute until exhaustion, with a fixed pedalling cadence between 60 and 80 rpm. The recovery phase included 2 min of active recovery at 20 W with a pedalling rate between 50 and 60 rpm, followed by 3 min of passive recovery. Measurements of gas exchanges (oxygen uptake: V̇O_2_, carbone dioxide production: V̇CO_2_, respiratory exchange ratio: RER, minute ventilation: V̇E, end-tidal pressure in oxygen and carbone dioxide: PETO_2_ and PETCO_2_) were performed every four respiratory cycles during testing with the following metabolic systems (Oxycon Pro, Jaeger, Germany and Cosmed Quark CPET, Cosmed, Italy), with data averaged every 15 seconds. Continuous electrocardiographic monitoring was conducted using the following ECG device (Case 12, St. Louis, USA and QuarkT12 ECG, Cosmed, Italy) and the rate of perceived exertion was assessed using the Borg scale (6–20) every 2 minutes during exercise. Patients were considered to have reached maximal effort when three of the following four criteria was met: (1) a plateau in V̇O_2_ (<150 mL/min during the last 30 seconds of exercise); (2) a respiratory exchange ratio greater than 1.10; (3) the inability to maintain a pedalling rate of 50 rpm; or (4) patient exhaustion due to fatigue or other clinical symptoms, including ECG or blood pressure abnormalities that necessitated stopping the exercise ^22, 34, 35^. The highest O_2_ uptake value (15s averaged) achieved during the exercise phase was considered the V̇O_2_peak. Peak power was defined as the highest power attained at the final fully completed exercise stage.

To compare cardiorespiratory fitness between women and men with CHD, the V̇O_2_peak value was normalized in % attained of the predicted value derived from the Wasserman equation (in % predicted) and to fat-free mass (V̇O_2_peak: mL/min/FFM kg) as previously published ^22, 34, 35^. The ΔV̇O_2_peak (mL/min/FFM kg) was calculated using the following formula: ΔV̇O_2_peak (mL/min/FFM kg) = V̇O_2_peak (mL/min/ FFM kg) post-training - V̇O_2_peak (mL/min/ FFM kg) pre-training. Oxygen uptake efficiency slope (OUES), the minute ventilation/carbon dioxide production slope (V̇E/V̇CO_2_ slope) and the ΔV̇O_2_/ΔWatts slope values were calculated according to published studies and recent recommendations ^22, 34–36^. Alveolar ventilation (V̇A: mL/min) was calculated using the following formula ^27, 35^:

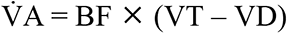

Where BF represents breathing frequency, and VT and VD refer to tidal and dead space volumes, expressed in mL, respectively. VD was estimated as previously suggested ^37^. The alveolar–arterial gradient (PAi-aO_2_, mmHg) was calculated using the following formula ^38^:

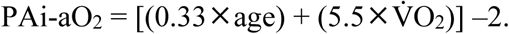

Cardiac hemodynamic parameters, such as Q̇c (L/min), CI (L/min/m²), and stroke volume (SV, mL), were measured on a beat-by-beat basis using a non-invasive impedance cardiography device (PhysioFlow Enduro®, Manatec, France) before and after the CR program ^34, 35^. These measurements provided reliable relative changes in cardiac hemodynamics between before and after CR. Subsequently, the data were averaged over 15 consecutive seconds. The arteriovenous oxygen difference C(a-v̅) O_2_, expressed in mLO_2_/100 mL, was calculated using the Fick principle ^35^:

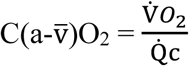

Exercise efficiency at peak exercise was defined as the gross efficiency of the external work (as a percentage) which was calculated using the following formula: ηW (%) = [external work (J ⁄ s) ⁄ metabolic work (J ⁄ s)]∈100. External and metabolic work were obtained from peak power output and V̇O_2_peak ^39^. Blood pressure was manually measured with a manual sphygmomanometer (767-Series, Welch Allyn Inc., USA) and used to estimate the value of mean arterial pressure (MAP, mmHg) as follows:

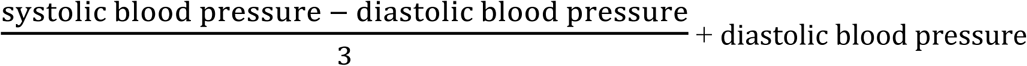

Therefore, CP, expressed in Watts, was calculated using the following formula ^40^:

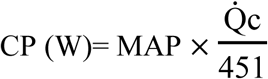

### Exercise training programs

The aerobic exercise training intervention included three distinct training modalities: low-volume high-intensity interval training (LV-HIIT), moderate-intensity continuous exercise training (MICET), and a combination of HIIT and MICET, as described in previous reports ^22, 34, 35^. All patients performed two to three supervised exercise training sessions per week on a bicycle ergometer. The total session duration was 33 to 45 min for LV-HIIT, 34 minutes for MICET, and 30 to 50 min for combined HIIT and MICET. Resistance training was performed following each aerobic session ^22, 34, 35^. Aerobic training load (training impulse: TRIMP) was calculated with the adapted model of Calvert et al. ^41^:

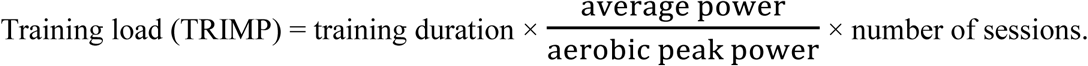

The training load is expressed in arbitrary units (A.U), training duration in minutes, and average and aerobic peak powers are in watts. Training load values and the program distribution for CHD patients stratified by responder status are presented in **Table 2**.

**Table 2.** Exercise training characteristics in women and men with coronary heart disease (CHD).

| **Variable** | **Women, n=28**<br>**(mean±SD)** | **Men, n=75**<br>**(mean±SD)** | ***p*-value** |
| --- | --- | --- | --- |
| Training duration (weeks) | 12.7±1.7 | 13.1±2.8 | 0.573 |
| Total sessions (n) | 32±5 | 33±4 | 0.137 |
| Weekly session (n) | 2.56±0.43 | 2.65±0.4 | 0.189 |
| Training adherence (%) | 92.8±11.0 | 95.7±10.8 | 0.374 |
| TRIMP (A.U) | 622.9±173.3 | 684.3±174.4 | 0.204 |
| HIIT (n, %) | 7 (25%) | 13 (17%) | 0.669 |
| MICET (n, %) | 3 (11) | 10 (14%) |  |
| Mixed (HIIT+MICET) (n, %) | 18 (64%) | 52 (69%) |  |
| Prevalence of responders |  |  | 0.826 |
| NR | 7 (27%) | 13 (19%) |  |
| LR | 4 (16%) | 13 (18%) |  |
| MR | 6 (23%) | 17 (24%) |  |
| HR | 9 (34%) | 28 (39%) |  |

A.U: arbitrary unit, NR: non responders, LR: low responders, MR: moderate responders, HR: high responders.

### Statistical Analysis

Continuous variables are presented as mean (standard deviation), while frequencies and percentages are presented for categorical variables. Statistical analyses were performed with JASP (v. 19.0, Netherlands) and Graphpad (v.10.2.2, USA). Baseline characteristics were compared between the women and men using either a t test or Mann-Whitney for continuous variables (depending on the distribution) and using a ꭕ^2^ for categorical variables. Mixed-model analysis (sex x time) was used to study the CPET parameters across time and between sex. Models with sex, time and sex x time interaction as independent variables were used. The sex x time interaction was the focus of the analysis as it tested the difference in the change (post-pre) between the 2 groups. Multiple comparison test was done using Šídák test to localize differences, with adjusted p-value (correction). To measure the magnitude of training in women and men with CHD, independently of sample size, the effect size (ES) was evaluated using the Hedge’s g. ES was characterized as trivial (d < 0.2), small (0.2 ≤ d < 0.5), moderate (0.5 ≤ d < 0.8) and large (d ≥ 0.8) ^42^. For the analysis of prevalence of responders, the ΔV̇O_2_peak (mL/min/FFM kg) was expressed in % change after the CR program. Then, the patients were classified as: non responders (NR: ΔV̇O_2_peak value ≤ 0%), low responders (LR: 0 < ΔV̇O_2_peak < 5%), moderate responders (MR: 5 ≤ ΔV̇O_2_peak < 10%) and high responders (HR: ΔV̇O_2_peak ≥ 10%) ^28, 35, 43, 44^. A significant difference between the groups was assessed at P<0.05.

## Results

### Clinical characteristics of the patients with CHD

A total of 153 patients with CHD (28 women, 18.3%) were eligible from the four prospective randomized trials. To remove potential baseline confounders before analysis, the sample of men with CHD was matched to that of women with CHD (n=28) on age, body mass index, and V̇O_2_peak (in % predicted). In the final analysis, we included a total of 28 women and 75 men with CHD (**Figure 1**: **flow chart**). The clinical characteristics are presented in **Table 1**. At baseline, age, BMI, V̇O_2_ peak (in % predicted) did not differ between women and men with CHD (p>0.05). Regarding peri-procedural characteristics, only percutaneous coronary intervention was higher in women than in men with CHD (p<0.05). Cardiovascular risk factors and baseline medications did not differ between women and men with CHD (p>0.05).

**Figure 1:**
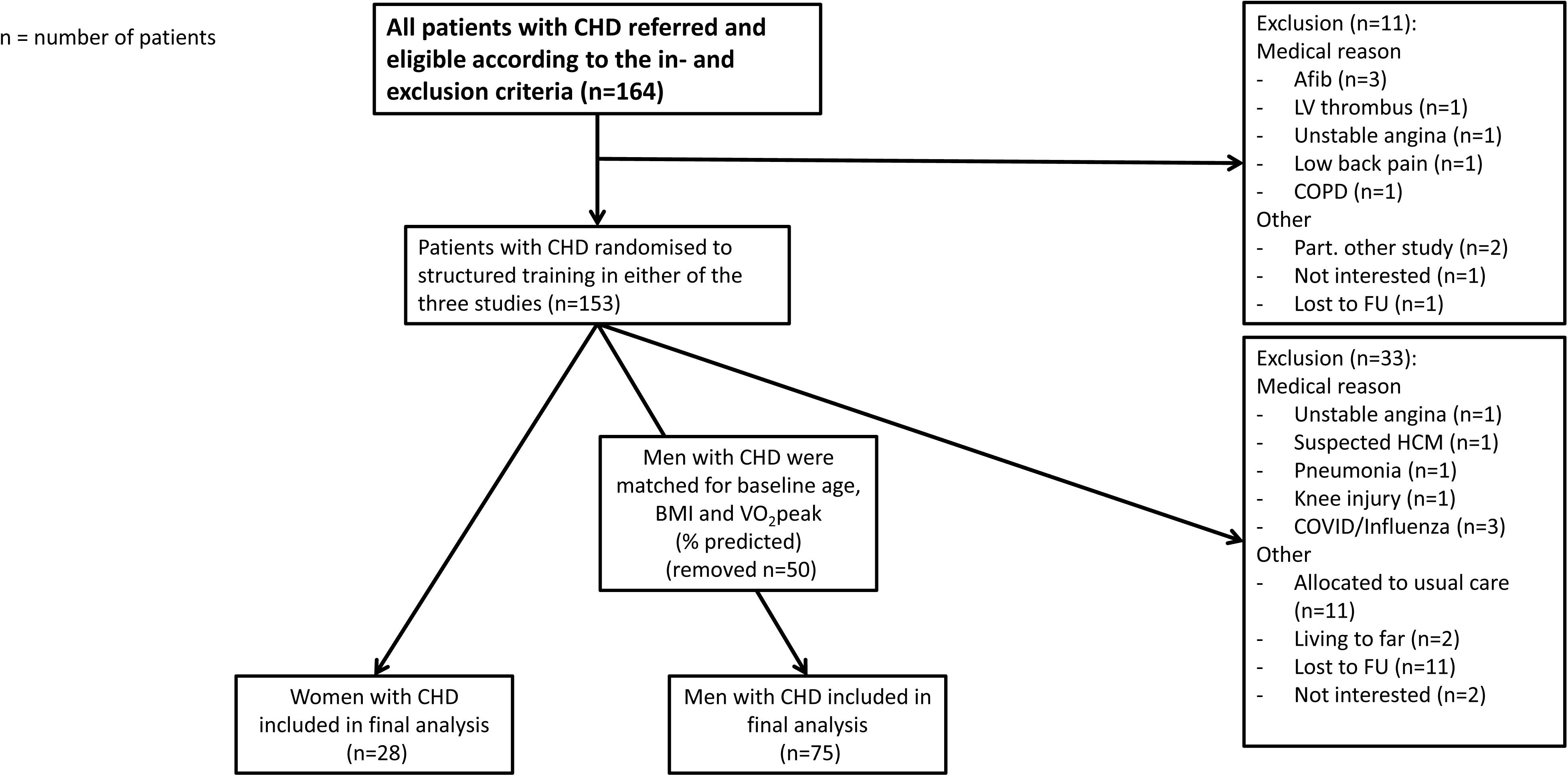
Flow chart of patient enrolment, matching, and final sample selection from four prospective randomized controlled trials. Legend: n represents the number of patients. CHD: coronary heart disease, Afib: atrial fibrillation, LV: left ventricular, COPD: chronic obstructive pulmonary disease, FU: follow-up, COVID: coronavirus disease, HCM: hypertrophic cardiomyopathy.

### Exercise training characteristics

Exercise training characteristics are given in **Table 2**. The training duration, total exercise sessions, exercise training adherence, exercise dose (training impulse: TRIMP), and exercise training composition (HIIT, MICET, mixed) did not differ between women and men with CHD (p>0.05).

### Determinants of V̇O_2_peak changes after exercise training

The prevalence of responders was also similar between women and men with CHD (p>0.05) (**Table 2**). The determinants of V̇O_2_peak in women and men with CHD are presented **in Table 3**. The changes in the determinants of V̇O_2_peak (expressed as a percentage) are depicted in **Figure 2** (radars). A group and time effect (p<0.0001) was shown for absolute and relative V̇O_2_peak (mL/min and mL/min/kg), indicating lower values for women vs. men with CHD, but a similar training effect (no significant interaction). A time effect (<0.0001) was shown for V̇O_2_peak normalized by FFM, suggesting a similar training effect between women and men with CHD.

**Figure 2:**
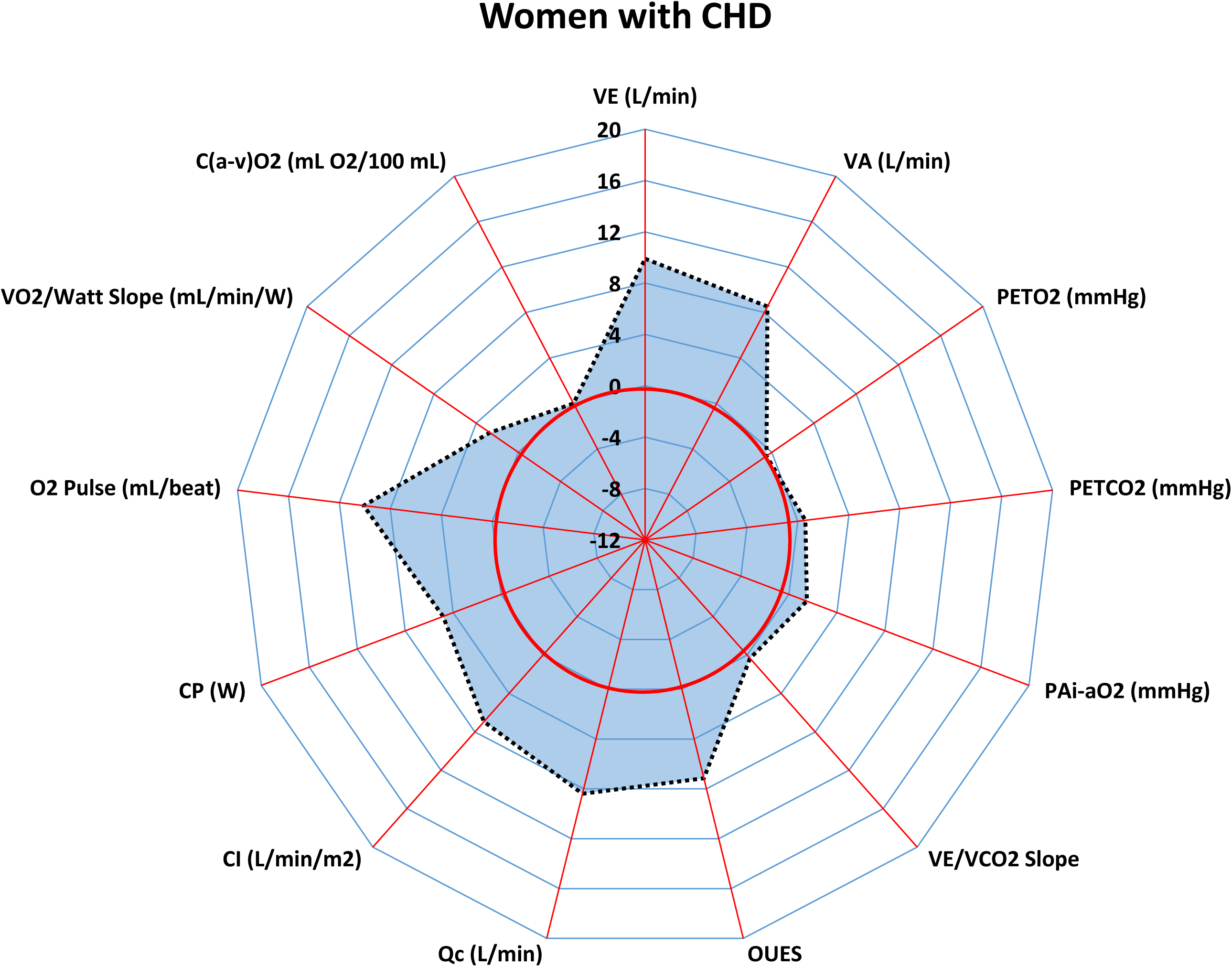
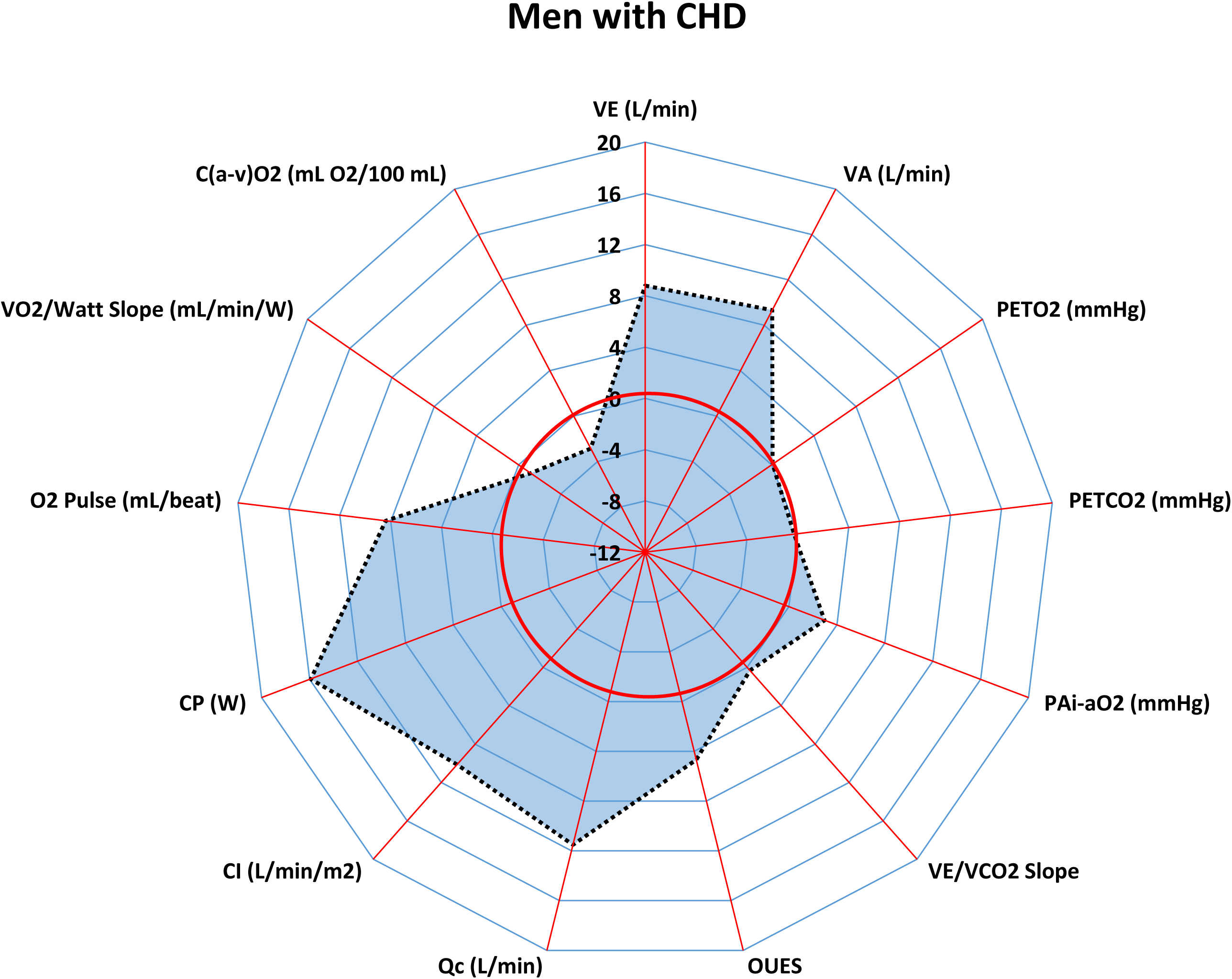
Percentage changes (%) in O_2_ transport chain determinants after CR in women (left panel) and men (right panel) with CHD. Values represent percentage changes ((Post-Pre)/Pre×100)) in cardiopulmonary exercise testing parameters. Abbreviations: V̇E: minute ventilation, V̇A: alveolar ventilation, PETO_2_, PETCO_2_: end-tidal pressures of O_2_ and CO_2_, PAi-aO_2_: alveolar-arterial O_2_ gradient, OUES, oxygen uptake efficiency slope; Q̇c, cardiac output; CI, cardiac index; CP, cardiac power; C(a-v)O₂, arteriovenous oxygen difference; ηW, gross muscular efficiency.

**Table 3.**
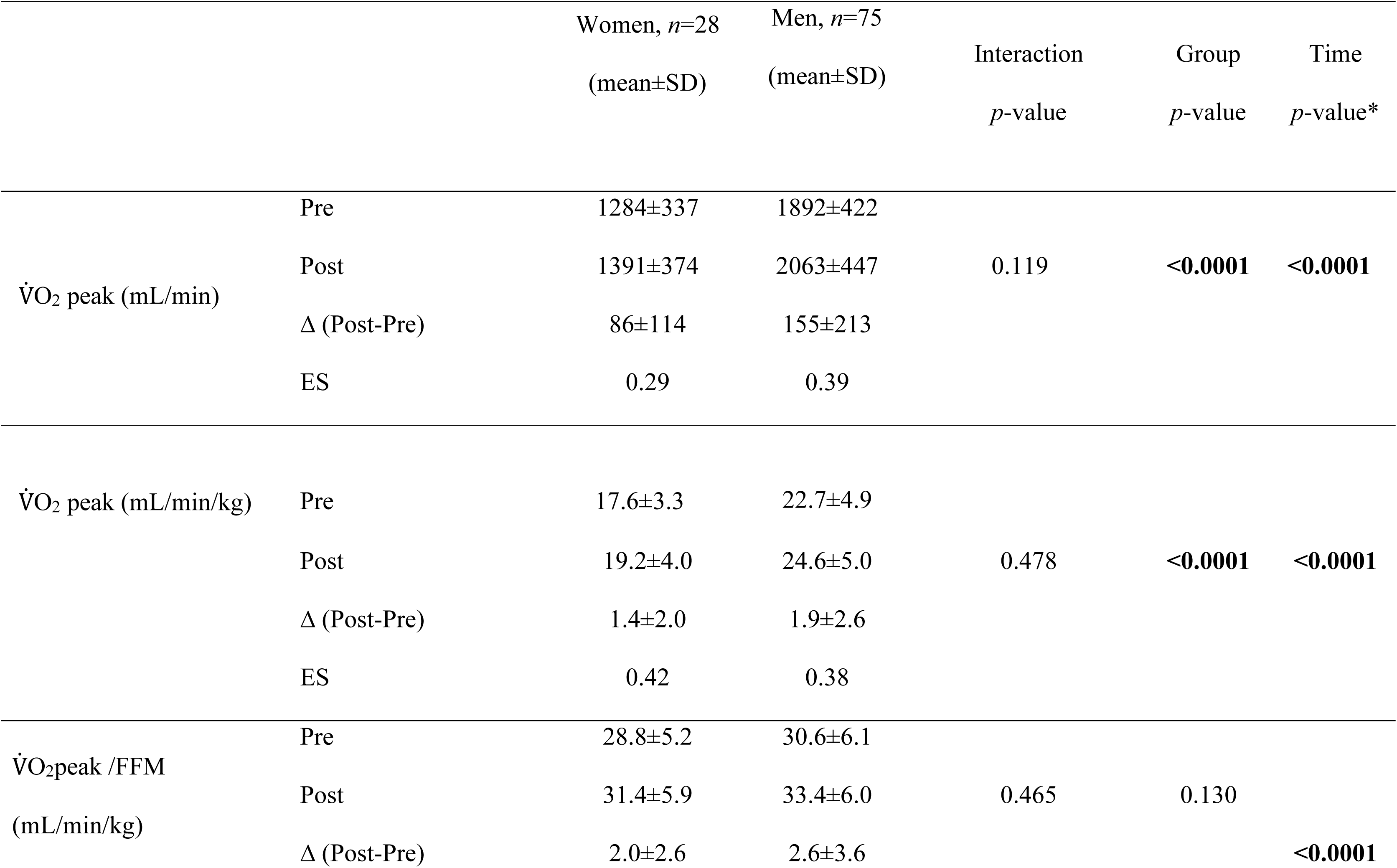
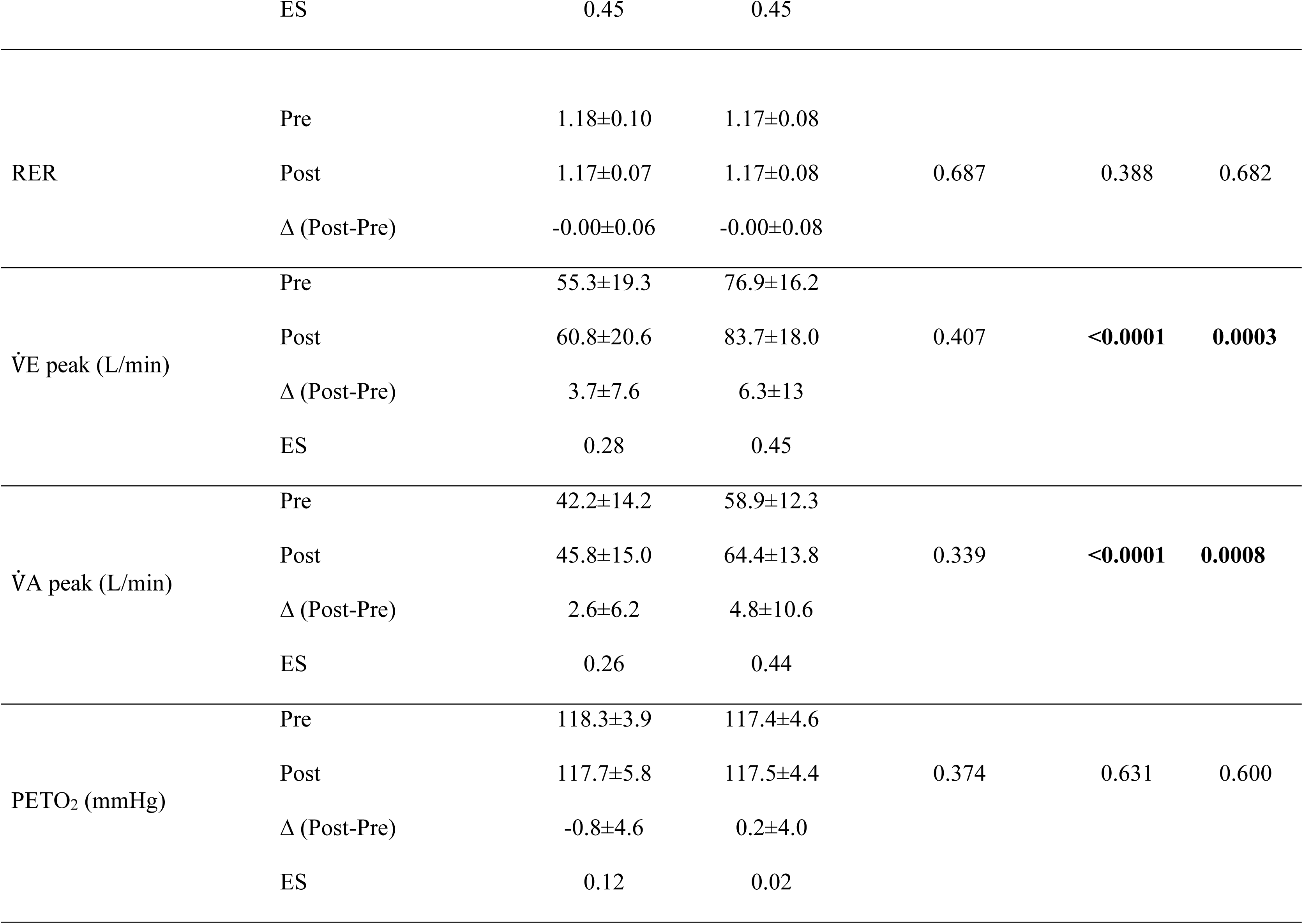
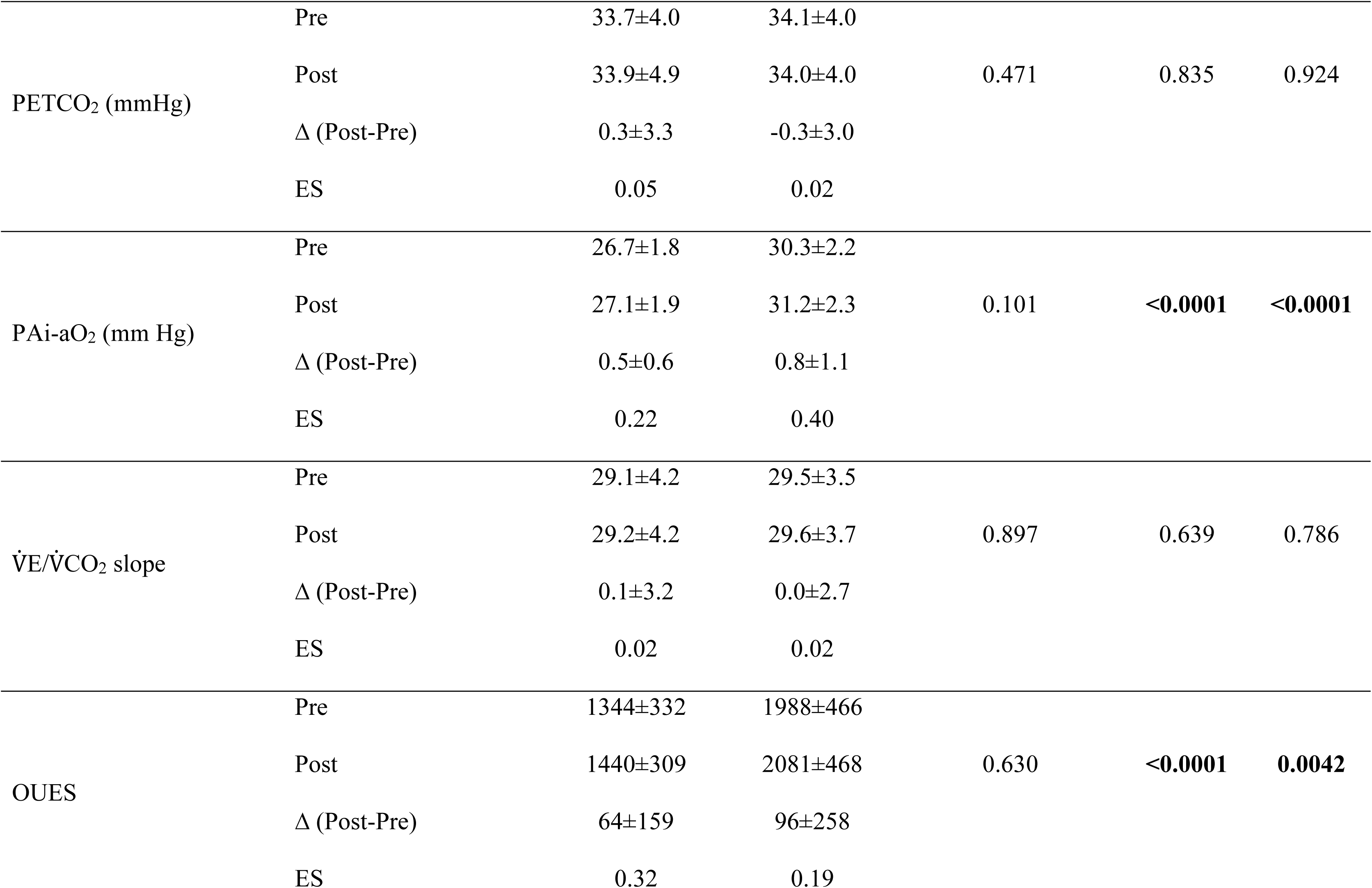
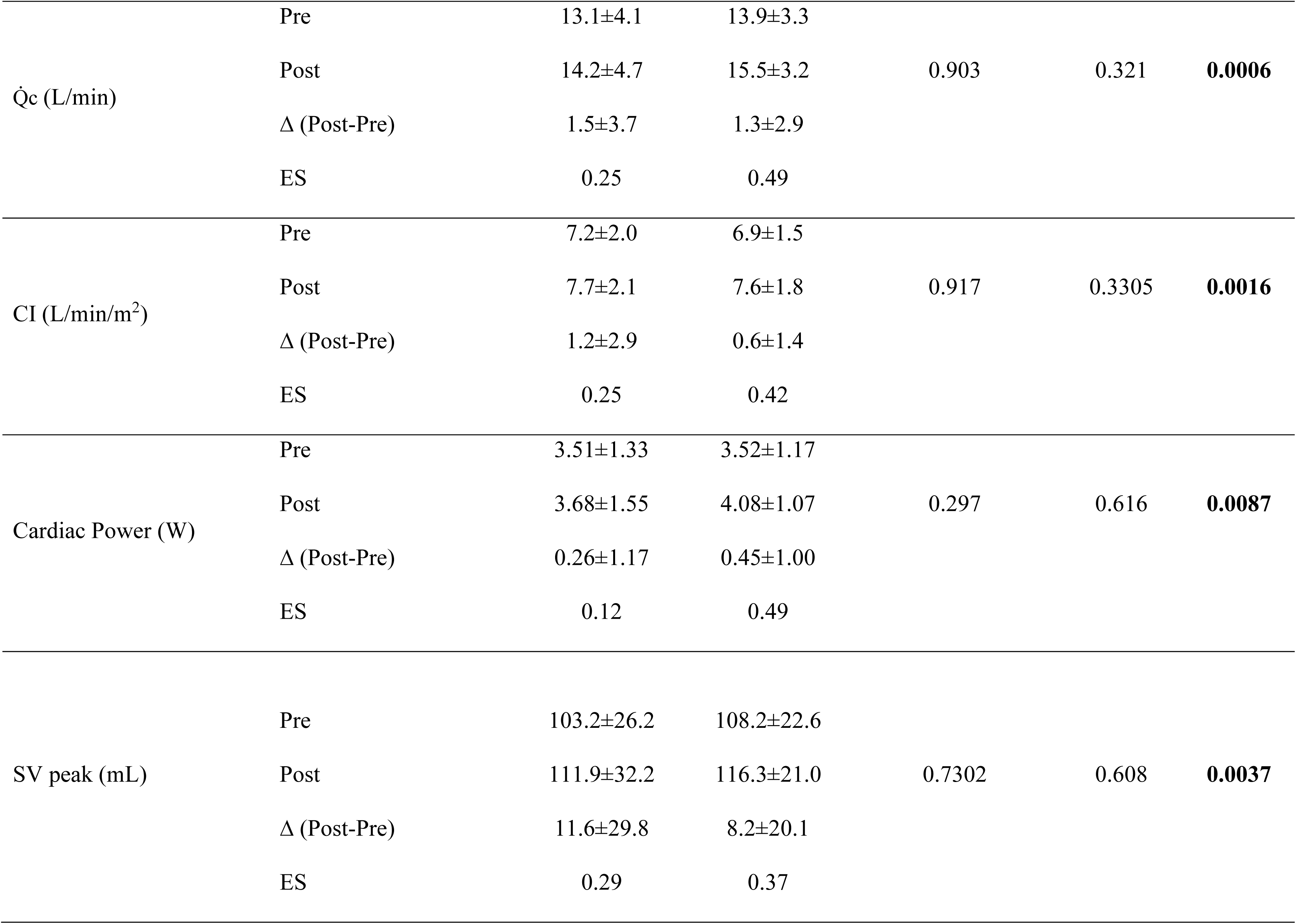
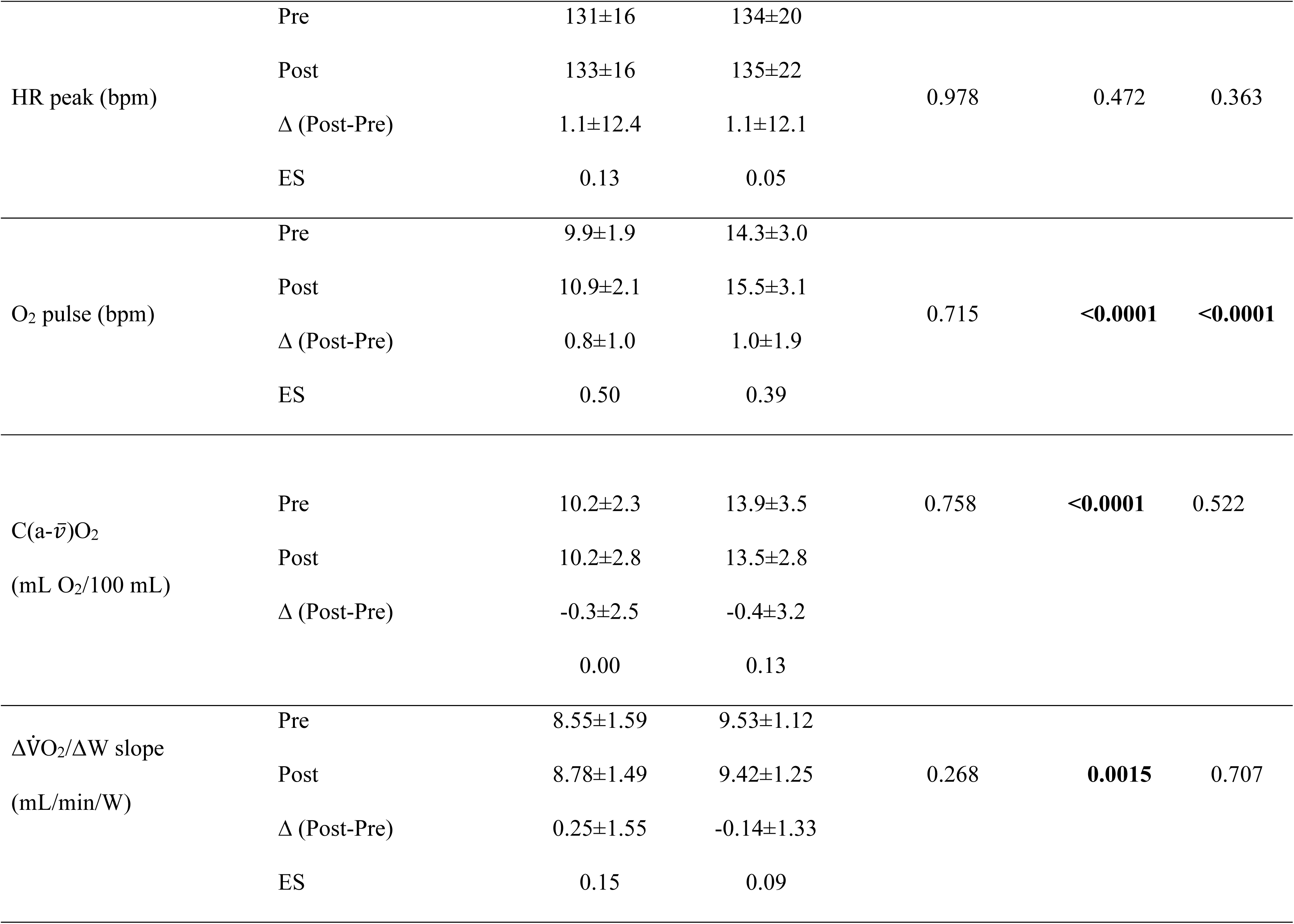
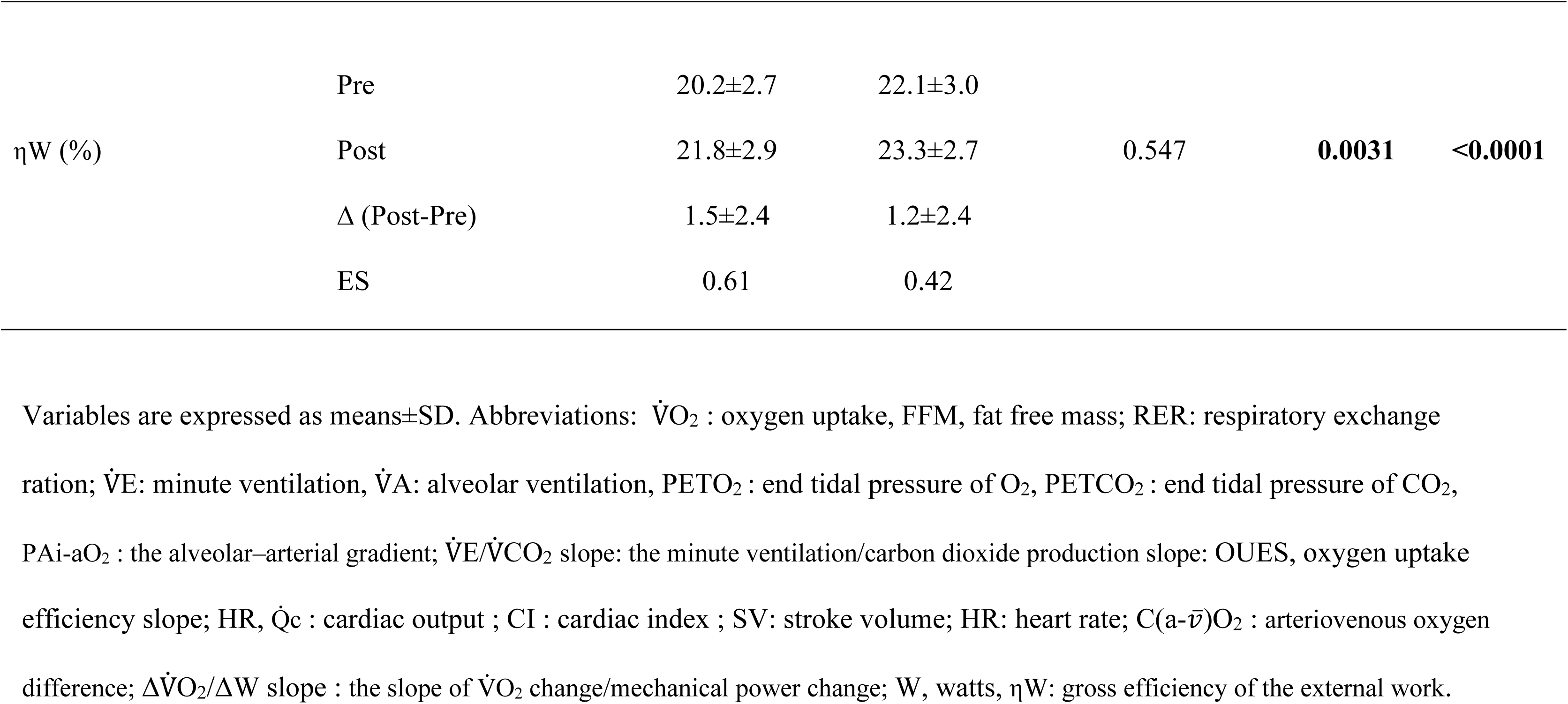
Determinants of V̇O_2_peak in women and men with coronary heart disease (CHD).

### Ventilatory and gas exchanges responses to exercise training

A group and time effect (p<0.0001) were observed for V̇E peak and V̇A peak, indicating lower values for women vs. men with CHD, but a similar training effect (no significant interaction). No significant group, time, or interaction differences were found for PETO_2_, PETCO_2_, and V̇E/V̇CO_2_ slope in women vs. men with CHD (p>0.05). A group and time effect (p<0.01) was observed for PAi-aO_2_ and OUES, with lower values for women than for men with CHD, but a similar training effect (no significant interaction).

### Cardiac and peripheral responses to exercise training

A significant time effect was found (p<0.01) for Q̇c, CI, CP and SV indicating a similar training effect between women vs. men with CHD on these variables. No significant group, time, or interaction differences were found for HR peak (p>0.05). A group and time effect (p<0.01) was observed for O_2_ pulse with lower values for women vs. men with CHD, but similar training effect (no significant interaction). A significant group effect was found (p<0.01) for C(a-v̅)O_2_ and the ΔV̇O_2_/ΔW slope, with lower values in women than in men with CHD. A group and time effect (p<0.01) was observed for gross efficiency of external work with lower values in women than in men with CHD, but similar training effect (no significant interaction).

## Discussion

The objectives of this study were to compare the ventilatory,circulatory and peripheral determinants of the underlying changes in V̇O_2_peak between women and men with CHD after exercise-based cardiac rehabilitation program. The main findings of the present study can be summarized as follows: 1) As expected, several absolute CPET values associated with pulmonary O_2_ convection (V̇E, V̇A), pulmonary O_2_ diffusion (PAi-aO_2_), ventilatory efficiency (OUES), circulatory convection (O_2_ pulse) and peripheral O_2_ diffusion (C(a-v̅)O_2_, ΔV̇O_2_/ΔW slope, ηW) were lower in women vs. men with CHD. 2) In addition to a similar V̇O_2_peak improvement, women with CHD showed a similar O_2_ transport phenotype adaptation as compared to men with CHD. Pulmonary O_2_ convection (V̇E, V̇A), pulmonary O_2_ diffusion (PAi-aO_2_), ventilatory efficiency (OUES), circulatory convection (Q̇c, CI, CP, O_2_ pulse) and peripheral muscular efficiency (ηW) were improved after CR to a similar extent in women and men with CHD (**Figure 2**). 3) With a similar exercise training dose and adherence, the prevalence of responder’s categories was not different between women and men with CHD. Contrary to our initial hypothesis, women with CHD have similar O_2_ transport phenotype adaptations vs. men with CHD after CR, namely with improved pulmonary O_2_ convection, circulatory convection, and peripheral muscular efficiency. From a clinical perspective, and due to the prognostic value of V̇O_2_peak improvement in women and men with CHD ^7, 8^, our results may have important implications for the referral and participation of women with CHD in CR programs. Indeed, Colbert et al. demonstrated that women completing a CR program experienced a greater relative reduction in mortality (HR 0.36) compared with men (HR 0.51), despite significantly lower referral rates (31% vs. 42%, p<0.0001) and completion rates (50% vs. 60%, p<0.0001) in women ^45^. These results suggest that the similar improvement in V̇O_2_peak and observed in our women with CHD might translate into equal, if not greater, prognostic benefits ^45^ which need to be confirmed in larger randomized controlled trials. This reinforces the urgent need to address the well-documented disparities in CR referral, access, and completion among women with CHD ^4^.

### V̇O_2_peak changes with exercise training

When expressed normalized by body mass, we found superior values for men (group effect) vs. women with CHD, in agreement with previous studies ^14, 15, 19, 22, 23^. Indeed, women generally have a higher fat mass percentage and lower FFM than men, as consistently reported in the literature, which might influence these results ^46, 47^. However, we found similar values and improvement of V̇O_2_peak normalized by FFM between women and men, with comparable effect sizes, underlying the importance of normalization of V̇O_2_peak with FFM ^22, 46, 47^. Since FFM represents the most metabolically active tissues implicated during V̇O_2_peak testing, this allows a more physiologically meaningful cardiorespiratory fitness comparison between sexes. In addition, women and men with CHD had a similar prevalence of V̇O_2_peak responder categories (from non to high response) after the exercise training program. Our results agree with previous studies regarding similar V̇O_2_peak improvement (directly measured by CPET, mostly expressed in mL/min/kg) between women and men with CHD after 8 weeks to 3 months of training ^19, 21–23, 25^. In contrast, other studies did show superior V̇O_2_peak improvement in men vs women with CHD (directly measured by CPET) after 3 to 9 months of training ^11, 14, 15, 17^. These conflicting results may partly be explained by the fact that our women and men with CHD were carefully matched for baseline age, body mass index and V̇O_2_peak (% predicted), minimizing the influence of key confounders on baseline and training responses. Another explanation may come from the FFM normalization and the similar exercise training dose, distribution and adherence (frequency, duration, number, and type of session) observed in our women and men with CHD. Our results replicate previous studies showing that when engaged in a CR program, women with CHD showed similar program adherence vs. their male counterparts ^17, 19, 24^. Other studies have reported a higher ^25^ or lower number of exercise sessions ^14^ in women vs. men with CHD.

### Ventilatory responses to exercise training

Regarding pulmonary gas exchange, our results suggest no significant changes in ventilation-perfusion matching or pulmonary diffusion capacity with training in both sexes as gas exchange indicators (PETO_2_, PETCO_2_, V̇E/V̇CO_2_ slope) remained unchanged after training, in agreement with previous studies in patients with CHD ^35^. Ventilatory efficiency (V̇E/V̇CO_2_ slope) was previously suggested to be similar in healthy older women than in men ^48, 49^, in agreement with our results, with a near-normal range value (mean: <30). Regarding gas exchange indicators (PETO_2_, PETCO_2_), previous studies suggest no sex difference ^49–51^, and pulmonary O_2_ diffusion during exercise was shown to be similar after adjusting for lung size in healthy women and men^50^. As expected, absolute peak V̇E and V̇A were significantly lower in women vs. men with CHD at baseline, reflecting well-known sex differences in lung size and pulmonary convective capacity ^22, 32, 46^. These differences could be largely explained by the smaller airway sizes and reduced tidal volumes found in women vs. men, previously impacting pulmonary function observed in both healthy individuals and patients with CHD^19, 22, 32, 46^. In addition, ventilatory convection (V̇E and V̇A) and ventilatory efficiency (OUES) showed similar improvements after exercise training in women and men with CHD, indicating that pulmonary adaptations (convection/efficiency) at peak effort were comparable between sexes. In women and men with CHD, previous studies are scarce and conflicting: one showed improvements in ventilation convection and efficiency (V̇E, OUES) after exercise training ^22^ and a second one showed no improvement in peak ventilation ^19^. For alveolar ventilation, this suggests a reduction in dead space in both sexes after training ^35, 52^, reinforced by the OUES improvement in our CHD groups. Improvement of OUES is consistent with previous studies showing that this ventilatory efficiency index ^22, 35, 53^ is sensitive to exercise training. This also suggests a greater efficiency of O_2_ extracted by the lungs rather than by muscle O_2_ extraction, since C(a-v̄)O_2_ remained unchanged after training in both sexes ^35, 54^.

### Cardiac and peripheral responses to exercise training

In an unexpected manner, the absolute values of peak cardiac output, stroke volume, and heart rate were not different among our women and men with CHD. This contrasts with what is generally reported in the literature with lower absolute peak cardiac output and stroke volume during exercise in patients with cardiovascular disease ^30, 31^, or in healthy older adults ^47, 55, 56^. The careful pairing of women and men with CHD based on factors such as age, BMI, and V̇O_2_ peak (% predicted) at the start may partly account for the reduced anthropometric differences that usually lead to sex-based variations in absolute cardiac hemodynamics. Additionally, the use of impedance cardiography for non-invasive cardiac hemodynamics assessment, which is recognized for its precision in monitoring relative changes rather than absolute values, might also have played a role in this finding ^35, 57^. In our study, the similar enhancement in peak cardiac output/power and stroke volume after exercise training, observed in our women and men with CHD, is significant and counterintuitive. It differs from the typical sex-specific cardiovascular adaptations seen in healthy older adults ^47, 56^, where men primarily enhance their V̇O_2_peak through central mechanisms ^58, 59^, such as increases in peak cardiac hemodynamics (Q̇c, SV at peak), while women mainly depend on improved peripheral oxygen extraction (C(a-v̄)O_2_), with minimal or no change in stroke volume. To our knowledge, no existing research has directly compared the effects of aerobic exercise training on peak cardiac hemodynamics (Q̇c, SV, CP) between women and men with CHD, making this study a unique contribution. Our results offer the first evidence that both women and men with CHD exhibit similar central cardiac adaptations from structured aerobic training. This conclusion cannot be drawn from previous studies on healthy older individuals due to the specific pathophysiological characteristics of coronary heart disease.

Regarding peripheral factors, women with CHD presented lower absolute values for muscular O_2_ extraction (C(a-v̄)O_2_) and efficiency parameters (ΔV̇O_2_/ΔW slope, ηW). The lower muscular efficiency parameters observed may be attributed to women’s reduced absolute peak workload and smaller muscle mass, rather than an inherent lack of mechanical efficiency. Lower absolute C(a-v̄)O_2_ values during exercise have been reported in older healthy women ^47, 56^ and women at risk for heart failure vs. their male counterparts ^31^. The reduced O_2_ extraction during exercise in women at risk for heart failure ^31^ may be due to both a lower arterial O_2_ content (resulting from decreased hemoglobin levels) and diminished muscle oxygen diffusion (DmO_2_), indicating a compromised O_2_ transport from the microvasculature to the mitochondria in skeletal muscles. Therefore, this could be speculative to apply these elements to our study because the C(a-v̅)O_2_ was calculated from Fick’s equation (V̇O_2_ and Q̇c). Indeed, major elements (blood gas analysis: arterial O_2_ content, hemoglobin, SaO_2_% and DmO_2)_ were unfortunately not measured in our study and remain to be performed in future studies to explore more deeply the exact muscular mechanism adaptations. After exercise training, there were no improvements in muscular O_2_ extraction during exercise and one muscular efficiency parameter (ΔV̇O_2_/ΔW) in both women and men with CHD. Some previous studies have demonstrated improvements in muscular O_2_ extraction in patients with CHD ^60, 61^, for the high responder’s CHD patients ^35^, but cardiac mechanisms remaining an important factor for V̇O_2_peak improvement particularly when exercise dose/intensity is increased ^26, 34^. Therefore, the clinical characteristics of CHD patients (i.e., age, CHD etiology, comorbidities), the intensity and duration of exercise training, as well as initial C(a-v̅)O_2_ close to normal values (10-14 mL O_2_/100 mL) might explain differences across studies. The lack of change in the ΔV̇O_2_/ΔW slope for both groups from pre- to post-training can be attributed to the nearly normal baseline values (9 mL/min/W), which offered little opportunity for enhancement, and this aligns with previous findings ^22, 35^. However, we observed a similar improvement of muscular external work efficiency (ηW) in both women and men with CHD. This enhancement indicates a greater capability of the musculoskeletal system to transform metabolic energy into external mechanical work, implying significant peripheral muscular adaptations in both sexes, even though C(a-v̄)O_2_ and the ΔV̇O_2_/ΔW slope remained unchanged. The improvement in mechanical efficiency in women with CHD holds clinical significance, as enhancing mechanical efficiency can lower the metabolic demands of everyday physical activities. This improvement could be especially advantageous for women with CHD, who often begin cardiac rehabilitation with reduced functional capacity ^30^.

### Study limitations

Initially, the recruitment process was conducted at a single institution, where there was a higher proportion of men with CHD participating in CR programs. Secondly, the estimation of alveolar diffusion was conducted using a specified formula; however, spirometry was not performed. Consequently, ventilatory reserve could not be accurately assessed. In addition, blood gases and haematological parameters were not assessed, and cardiac hemodynamic responses were evaluated using a non-invasive method (impedance cardiography: ICG). Nonetheless, a rigorous protocol was implemented, which included a smoothing process and the exclusion of low-quality ICG data. The Fick equation might be speculative in accounting for all the findings, especially when C(a-v̅)O_2_ was determined with the Fick’s equation and calculated C(a-v̅)O_2_ value may be quite higher. Although the absolute values from ICG have been debated, this methods is validated for detecting relative changes, which was the focus of the present study ^57^.

## Conclusion

This study is the first to demonstrate that both women and men with CHD, when matched for initial age, body mass index, and cardiorespiratory fitness, exhibit comparable adaptations in the O_2_ transport phenotype after participating in a structured exercise-based cardiac rehabilitation program. Contrary to our initial hypothesis, women with CHD did not exhibit a predominantly peripheral adaptation pattern, as is typically observed in healthy older women. Instead, they showed similar enhancements across key stages of the O_2_ transport chain, including pulmonary O_2_ convection (V̇E, V̇A), ventilatory efficiency (OUES), circulatory convection (cardiac index and power, SV), and peripheral gross muscular efficiency (ηW). Additionally, the prevalence of V̇O_2_peak responder categories was comparable between sexes. The results indicate that the pathophysiological environment of CHD, when combined with complete coronary revascularization, optimal medical treatment, and a supervised exercise regimen with good adherence, reduces the typically observed sex-specific cardiovascular adaptation pattern in healthy older adults. The similar enhancement in V̇O_2_peak (when adjusted for fat-free mass) and in gross mechanical efficiency (ηW) in both sexes underscores the significance of considering body composition when evaluating training responses between women and men with CHD. Future studies, including more women with CHD, should investigate whether longer training programs or higher exercise doses can further differentiate O_2_ transport chain adaptations between sexes. Blood gas analysis in women and men with CHD will help to document more accurately the phenotypic adaptations for pulmonary O_2_ diffusion, hematological parameters of O_2_ transport, arterial O_2_ content, and muscle O_2_ diffusive conductance, providing deeper mechanistic insights. These results strengthen the rationale for improving CR referral, access, and adherence among women with CHD, and for using body-composition-adjusted fitness outcomes to characterize training responses across sexes, while future studies should determine whether these equivalent O₂ transport adaptations translate into comparable or superior long-term reductions in cardiovascular events and mortality in women.

## Authors contributions

M.G. participated in study conception, data analysis, statistics and draft of the article.

A. F. B. participated in data analysis, revision, statistics, and revision of the article.

PM.L. participated in the draft of the manuscript, data analysis and revision, statistics, revision of the article.

LD.T. participated in data analysis and revision, statistics, and revision of the article.

J.I-G: A.G. participated in data analysis, revision, and statistics, and revision of the article.

A. M. B. participated in data analysis and revision, statistics, and revision of the article. P-O.M participated in data analysis and revision, statistics, and revision of the article.

D.V participated in data analysis and revision, statistics, and revision of the article.

M.K participated in data analysis and revision, statistics, and revision of the article.

A.G. participated in data analysis, revision, and statistics, and revision of the article.

J.L. contributed to data collection, analysis, and study coordination.

M.J. participated in study conception, data analysis, and revision of the article, study coordination and financial study aspects.

A. N. participated in study conception, revision of the article, and study coordination.

L.B. participated in the revision of the article, study coordination, and financial study aspects.

## Data Availability

All data produced in the present study are available upon reasonable request to the authors

